# Assessing the potential impact of immunity waning on the dynamics of COVID-19: an endemic model of COVID-19

**DOI:** 10.1101/2021.10.23.21265421

**Authors:** Musa Rabiu, Sarafa A. Iyaniwura

**Affiliations:** School of Mathematics, Statistics & Computer Science, University of KwaZulu-Natal, Durban, South Africa; Department of Mathematics and Institute of Applied Mathematics, University of British Columbia, Vancouver, BC, Canada

**Keywords:** COVID-19, endemic model, backward bifurcation, non-pharmaceutical intervention, vaccination, immunity waning

## Abstract

We developed an endemic model of COVID-19 to assess the impact of vaccination and immunity waning on the dynamics of the disease. Our model exhibits the phenomenon of backward bifurcation and bi-stability, where a stable disease-free equilibrium co-exists with a stable endemic equilibrium. The epidemiological implication of this is that the control reproduction number being less than unity is no longer sufficient to guarantee disease eradication. We showed that this phenomenon could be eliminated by either increasing the vaccine efficacy or by reducing the disease transmission rate (adhering to non-pharmaceutical interventions). Furthermore, we numerically investigated the impacts of vaccination and waning of both vaccine-induced immunity and post-recovery immunity on the disease dynamics. Our simulation results show that the waning of vaccine-induced immunity has more effect on the disease dynamics relative to post-recovery immunity waning, and suggests that more emphasis should be on reducing the waning of vaccine-induced immunity to eradicate COVID-19.

## 1 Introduction

The Severe Acute Respiratory Syndrome Coronavirus (SARS-CoV) that infected millions of people around the world in 2002 and the Middle East Respiratory Syndrome Coronavirus (MERS-CoV) in 2012 are responsible for many cases of the common cold (Flu) in recent years. In mid-December 2019, twenty individuals suffering from pneumonia and seven other critically ill patients were admitted to the Wuhan Municipal Health Commission (WHC). The majority of them had prolonged exposure to animals such as snakes, poultry, bats etc., at the Huanan Seafood Market in Wuhan [7]. After extensive research by the World Health Organization (WHO) [37], these individuals were believed to be infected by a strange virus that was later named the Severe Acute Respiratory Syndrome Coronavirus 2 (SARS-CoV-2) by the WHO on February 11, 2020 [63]. The disease caused by the virus was also named COVID-19 on the same day [63]. As at August 29, 2021, there has been over 216,789,388 confirmed cases of COVID-19 globally, with 4,508,379 confirmed deaths and over 193,721,846 recovered individuals [65].

After infection, the corona-virus attacks the body cells and replicate quickly resulting in a severe cases of COVID-19 [48, 55]. According to the medical practitioners, COVID-19 victims experience mild or moderate respiratory problems and difficulty in breathing. Other symptoms include but not limited to tiredness, flu, cough, fever and shortness of breath [36, 64]. Infected individuals go through the incubation period of 2-14 days for developing symptoms, while some do not show symptoms at all during their infection (asymptomatic individuals) [34, 35]. It is believed that asymptomatic individuals account for more than 80% of the community transmissions [68]. The incubation period is generally referred to as asymptomatic stage since the infected person can still successfully transmit the disease around the community through effective contact [34, 35, 68]. The individuals in this category are generally encouraged by WHO to self-isolate for a minimum of 14 days and be in constant contact with the COVID-19 regulatory line in their community to forestall further transmission. People with severe illness, difficulty in breathing and those with underlying medical conditions are encouraged to visit the hospital with immediate alacrity [65]. According to the virologists, corona-virus is a single-stranded RNA virus while HIV is a retrovirus (ie. it has the ability to carry single-stranded RNA as its genetic material instead of double-stranded DNA carried by human cells) [16]. The implication of this is that the treatment used in treating diseases with RNA virus can also be used for COVID-19 treatment [17].

In an effort to curb the transmission of COVID-19, many countries and regions around the world implemented the non-pharmaceutical interventions (NPIs) such as physical distancing, wearing of face mask, quarantine, closure of schools/businesses, travel restriction, among others [12, 14, 22, 44, 45, 57]. The implementation of some of these NPIs have both physical and mental implications on the society. Some of these include loss of jobs, loneliness, lack of access to adequate medical treatment, economic meltdown, etc. [6, 15, 25, 41, 45, 50, 54]. The availability of COVID-19 vaccines has represented a unique opportunity in the fight against COVID-19. In addition to reducing disease transmission and burden, widespread vaccination will also allow countries to safely lift NPIs and promote the gradual recovery from the pandemic. Vaccination has been reported as one of the best development in the public health sector since around 1900 [23, 30, 31, 32]. It has helped in the eradication of several infectious diseases such as polio, measles, rubella, smallpox etc [19]. Despite the public health benefits derived from vaccination, there are still a lot of concerns about the need, efficacy and safety of many available vaccines [23, 30, 31, 32].

South Africa is one of the first African countries to receive the COVID-19 vaccine. They received their first shipment which contains a million doses of the AstraZeneca/Oxford COVID-19 vaccine, on February 1, 2020 [60, 62]. A few days later, the vaccination process was put on hold over concerns on the efficacy of the vaccine against the B.1.351 variant of SARS-CoV-2, which was the dominant strain of the virus in the country [40, 42]. The Johnson and Johnson COVID-19 vaccine, with an initial 80,000 doses [43], was rolled out on the February 17, 2021. This vaccine is observed to be more effective against the B.1.351 strain [40, 42].

Several mathematical models have been developed and analyzed to study the transmission dynamics of COVID-19 [2, 4, 26, 27, 33, 53, 39, 46, 66, 70]. Bugalia et al. [4] developed a COVID-19 model using an endemic framework considering the roles of intervention strategies such as lockdown, hospitalization and quarantine. They computed the reproduction number and calculated both the disease-free and endemic equilibriums of their model. They also established the epidemiological significance of the equilibrium stability, transcritical bifurcation and boundedness of solution. In order to validate their system of differential equations representing the COVID-19 dynamics, they fitted the cumulative and new daily cases in India after which they estimated the model parameters and predicted the near future scenario of the disease. The global sensitivity analysis was also established to observe the impact of different parameters of *R*_*o*_. They also investigated the dynamics of disease in respect to partial lock-down, complete lock-down and no lock-down. Their analyses conclude that if there is partial or no lockdown case, then endemic level would be unbelievably high and that India may experience more that six million infections if care is not taken.

The effect of an imperfect vaccine in reducing COVID-19 transmission and eventually eradicating the disease in the United State of America was examined in [24]. Their mathematical model provides an estimate for the minimum number of susceptible individuals needed to be vaccinated to obtain vaccine-induced herd immunity. In addition, they investigated the epidemiological consequence of the herd immunity threshold and showed that the disease can be eliminated if the herd immunity threshold is achieved. They simulated their model using COVID-19 parameters related to states like Florida and New York, and estimated the herd immunity threshold for these states. The impact of government actions and individual reactions on the corona-virus dynamics in South Africa was assessed using a mathematical model by [47]. Their model examines the effects of individual behavioral reaction and non-pharmaceutical interventions such as quarantine and lockdown on the dynamics of COVID-19 in South Africa. Based on the South African officially published COVID-19 data for the period of March 2020 to early May 2020. They discovered that a detection rate of at least 0.5 per day may lead to a significant reduction in the total number of infected cases. They also observed that in the absence of intervention strategies, the peak number of cases could be attained earlier than expected which can also be delayed if the interventions are implemented. The importance of having a COVID-19 vaccine that is effect in older individuals was emphasized in [53].

Johnson and Bruce [29] proposed a framework for modeling fear of infection and frustration with social distancing during COVID-19 epidemic. They established that the SEIR behavior-perception model has three principal modes of qualitative behavior: no outbreak, controlled outbreak, and uncontrolled outbreak. They fitted their model to the cumulative cases of COVID-19 and mortality data for different regions, and showed that their model can produce and sustain waves of infections. Their results show that the region with significant decline after wave of infection may likely survive the second wave, while those with moderate success in controlling their initial COVID-19 outbreak are most likely to experience substantial second waves or devastating outbreaks.

In this work, we develop and analyze a novel endemic model of COVID-19 that incorporates vaccination as a pharmaceutical intervention strategy (PIS). This model considers the waning of both vaccine-induced immunity and post-recovery immunity. We derive the control reproduction number for our model and study the disease-free and endemic dynamics of the model with a focus on the South African COVID-19 scenario. Our goal is to use this model to study the effect of immunity waning on both the control reproduction number and the disease dynamics.

The rest of the paper is structured as follows. In Section 2, we present the model formulation and state the assumptions on the model. Section 3 entails the model analyses. In the first part, we discuss the non-negativity of solution and the invariant region, while in the second part we establish the bifurcation analysis and discuss its biological interpretations. In Section 4, we present numerical computations of the control reproduction number derived in Section 3, in terms of the vaccination and immunity waning parameters. Also in this section, we study the effects of vaccination and immunity waning of the dynamics of the disease. A brief discussion and future work conclude the paper in Section 5.

## 2 Mathematical model

We develop a six compartment endemic model of COVID-19 to study the effect of immunity waning on the dynamics of the disease. Our model incorporates vaccination as a pharmaceutical intervention strategy and assumes that the immunity provided by the vaccine (vaccine-induced immunity) and immunity due to infection (post-recovery immunity) wane over time [18, 8, 56, 21]. The implementation of vaccination in our model is such that it reduces the chances of susceptible individuals acquiring the infection. The susceptible compartment is divided into two: unvaccinated susceptible population (*S*) and vaccinated susceptible population (*S*_*v*_). We also divided the infected population into three: exposed (not infectious) (*E*_1_), pre-symptomatic (infectious without symptoms) (*E*_2_) and symptomatic infectious (*I*). The recovered population is denoted by *R*. We assume that recovered individuals do not have permanent immunity to the disease, they become susceptible again at some rate (post-recovery immunity waning). The vaccine and post-recovery immunity waning assumptions in the model can also be interpreted as vaccinated and recovered individuals becoming susceptible to another variant of the virus. Table 1 gives the descriptions of the variables in our model.

**Table 1:** Model variables and descriptions. Our model assumes that individuals in the exposed compartment (E_1_) are not infectious, while those in the pre-symptomatic compartment (E_2_) are infectious but do not show symptoms.

| Variable | Description |
| --- | --- |
| $S$ | Unvaccinated susceptible population |
| $S_v$ | Vaccinated susceptible population |
| $E_1$ | Exposed population |
| $E_2$ | Pre-symptomatic infectious population |
| $I$ | Symptomatic infectious population |
| $R$ | Recovered population |

Figure 1 shows a schematic diagram of the model, where the black and green arrows indicate the direction of the flow of individuals between the compartments at the rates indicated beside the arrows. The green arrows specifically show the flow of individuals due to immunity waning. The red arrows show natural death and death due to COVID-19, while the blue arrow shows birth and immigration into the susceptible population. Our model assumes that immigration into the population only happens in the susceptible population and individuals do not leave the population through emigration. The transmission rates for the pre-symptomatic and symptomatic individuals are respectively denoted by *β*_1_ and *β*_2_. Using these two rates, we define our force of infection, *λ* as

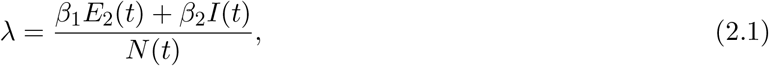

where *N* (*t*) = *S*(*t*) + *S*_*v*_(*t*) + *E*_1_(*t*) + *E*_2_(*t*) + *I*(*t*) + *R*(*t*) denotes the total population at any time *t*. The differential equations of our model are given by

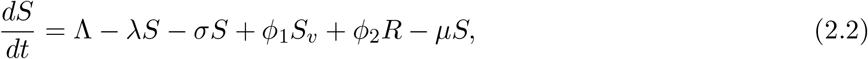

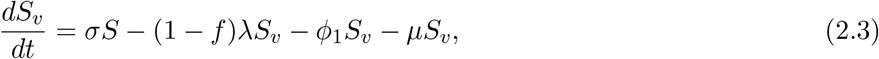

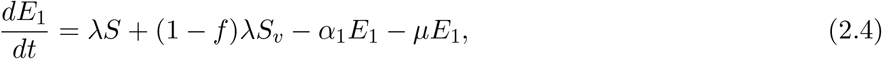

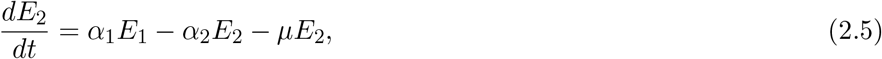

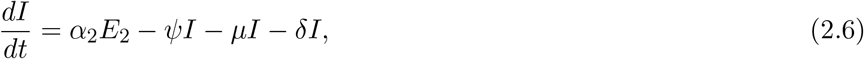

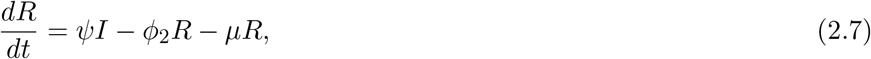

under the following initial conditions

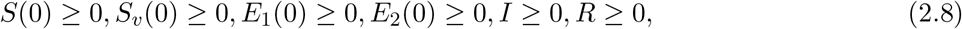

**Figure 1:**
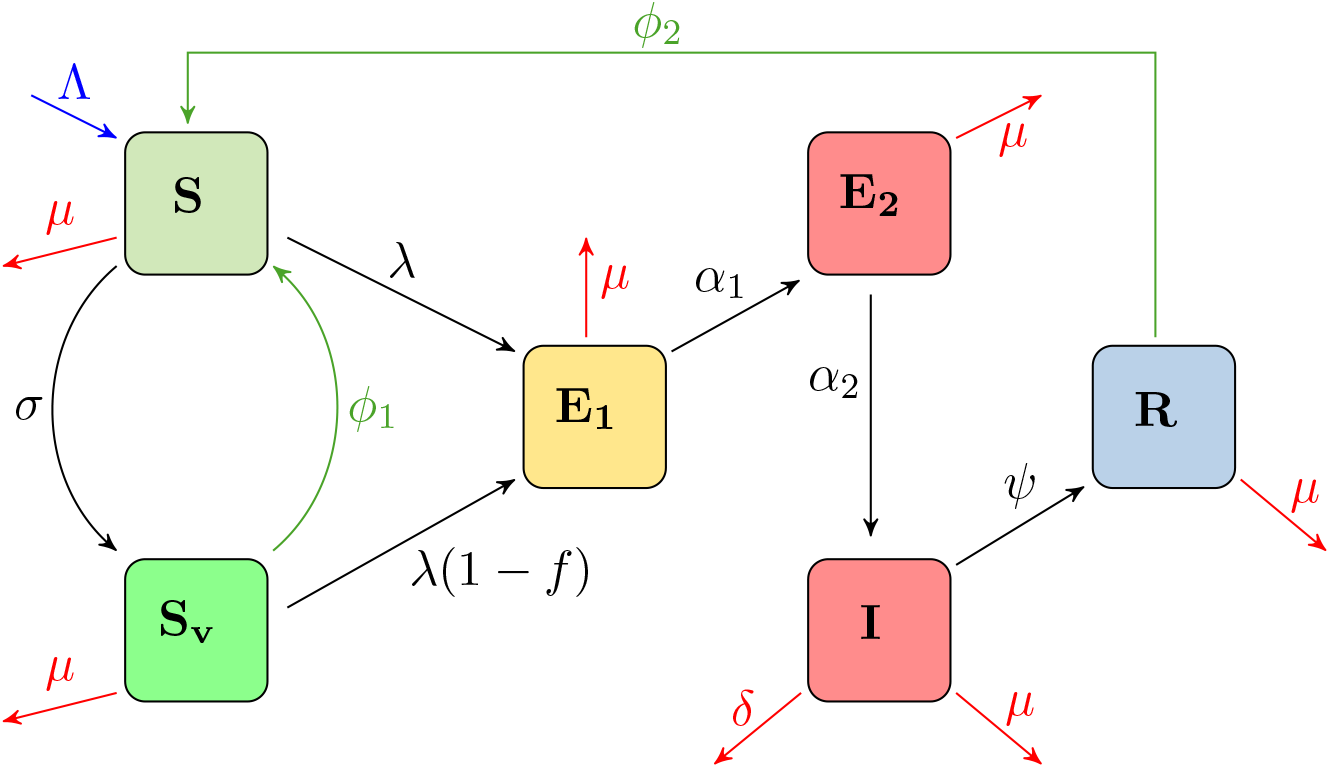
Model schematic. Compartments are as follows: Unvaccinated susceptible (S); vaccinated susceptible (S_v_); exposed (E_1_); pre-symptomatic infectious (E_2_); symptomatic infectious (I) and recovered (R). Black solid arrows show the flow of individuals through the compartments of the model from susceptible to recovered. The green arrows show the return of individuals to the susceptible compartment due to immunity waning. The blue arrow indicates birth and immigration into the susceptible compartment, while the red arrows show natural death and death due to COVID-19. The rates of transitioning from one compartment to the other are shown beside the arrows (see equations (2.2)-(2.7) for more details).

where *σ* is the vaccination rate and 0 ≤ *f* ≤ 1 is the vaccine efficacy. Here, *f* = 0 implies that the vaccine does not provide any protection from acquiring the disease, while *f* = 1 implies that it is 100% effective in preventing new infection. The vaccine and post-recovery immunity waning rates are *ϕ*_1_ and *ϕ*_2_, respectively. The natural death rate is denoted by *µ*, while the rates of transitioning from *E*_1_ to *E*_2_ and *E*_2_ to *I* are denoted by *α*_1_ and *α*_2_, respectively. Symptomatic infectious individuals recover from the disease at the rate *ψ* and die as a result of COVID-19 related complications at the rate *δ*.

In Table 2, we present the model parameters, their descriptions and values. The birth and immigration rate Λ, and the death rates *µ* and *δ* are related to South African demography. These parameters were obtained from the official Republic of South Africa websites. The vaccine and post-recovery immunity waning parameters are varied throughout this paper. The exact values used in each scenario will be specified in the figure caption.

**Table 2:** Model parameters, descriptions, and values. The birth and immigration rate Λ, and the death rates µ and δ are based on South African demography, and were obtained from the official Republic of South Africa government websites [1, 61].

| Parameter | Description | Value | Reference |
| --- | --- | --- | --- |
| $\Lambda$ | Birth and immigration rate | 2795 day<sup>-1</sup> | [1] |
| $\beta_1$ | Transmission rate for pre-symptomatic population | 0.2189 | [47, 67] |
| $\beta_2$ | Transmission rate for symptomatic population | 0.3521 | [47, 67] |
| $\lambda$ | Force of infection | Computed | See (2.1) |
| $\sigma$ | Vaccination rate | Varied | |
| $f$ | Vaccine efficacy | Varied | |
| $\phi_1$ | Vaccine immunity waning rate | Varied | |
| $\phi_2$ | Post-recovery waning rate | Varied | |
| $\mu$ | Natural death or emigration rate | 0.0070 | [1] |
| $\alpha_1$ | Rate of transitioning from $E_1$ to $E_2$ | 1/5 day<sup>-1</sup> | [47, 33, 71] |
| $\alpha_2$ | Rate of transitioning from $E_2$ to $I$ | 1/2 day<sup>-1</sup> | [38, 47, 58, 16] |
| $\psi$ | Recovery rate | 1/7 day<sup>-1</sup> | [47, 58, 16] |
| $\delta$ | COVID-19 induced death rate | 0.0014 | [69, 49, 28, 61] |

## 3 Model analysis

In this section, we analyze our ODE model (2.2)-(2.7) and show the non-negativity of its solution and that of the invariant region that describes the solution. We also analyze the linear stability of disease-free equilibrium and the endemic equilibrium.

### 3.1 Non-negativity of Solution and the Invariant Region

Here, we establish the non-negativity of the solutions of the ODE model (2.2)-(2.7) as well as the invariant region that describes these solutions.

#### Lemma 1.

*Let ϒ represents the closed set defined as*

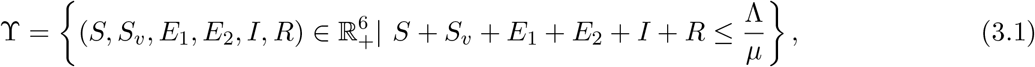

*then* *equation* (3.1) *is positively invariant and attracting with regards to* (2.2)*-*(2.7).

*Proof*. Using (2.8) as the initial conditions of the ODE system (2.2)-(2.7), from (2.2), we have

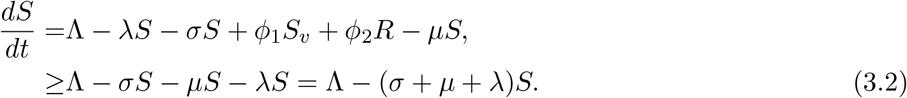

Observe that equation (3.2) is a first-order linear differential equation. The integrating factor is chosen as

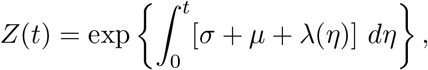

where *λ*(*η*) ≡ *λ*(*S, S*_*v*_, *E*_1_, *E*_2_, *I, R*) so that the solution to (3.2) with equality is

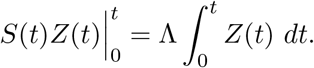

Evaluating the equation above and using the fact that *S*(0) ≥ 0, we have

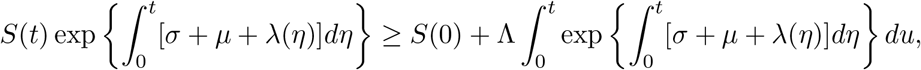

which implies that

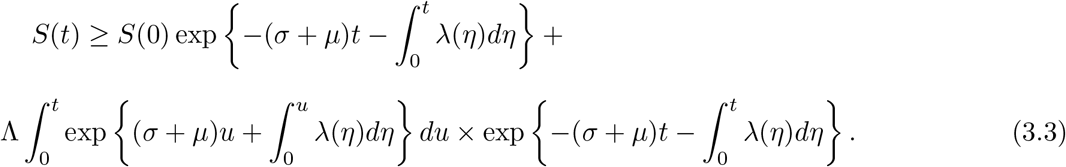

Since *S*(0) is a non-negative constant (see (2.8)), (3.3) ensures that the solution *S*(*t*) is non-negative.

Following the same approach, the non-negativity of *S*_*v*_, *E*_1_, *E*_2_, *I* and *R* can be established provided that the initial conditions exist for all time *t* > 0.

More so, summing up equations (2.2)-(2.7) gives

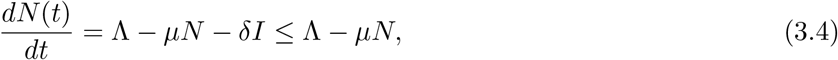

whose solution is given by

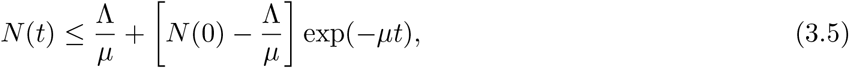

where exp(*µt*) is the integrating factor. Taking the limit of both sides as *t* → ∞ gives

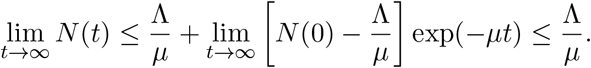

This means that the expression in (3.5) is bounded above by 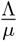 in the domain described in Lemma 3.1. Hence, the ODE system (2.2)-(2.7) is well-posed epidemically and meaningful mathematically since all the state variables are non-negative for all *t* > 0. Thus, we proceed to analyze the model in ϒ. This completes the proof. □

### 3.2 Local Stability of Disease-free equilibrium (DFE)

The disease-free equilibrium of (2.2)-(2.7) is given by

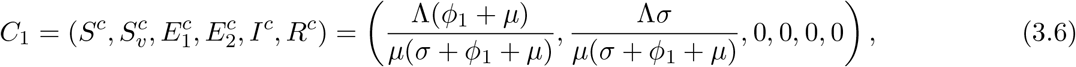

where 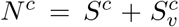. We calculate the control reproduction number using the next generation matrix operator [13, 59] on (2.2)-(2.7) as used by [46, 51]. The matrix for the rates of new infections ℱ and that of transfer of infected individuals, 𝒱 at the DFE are given by

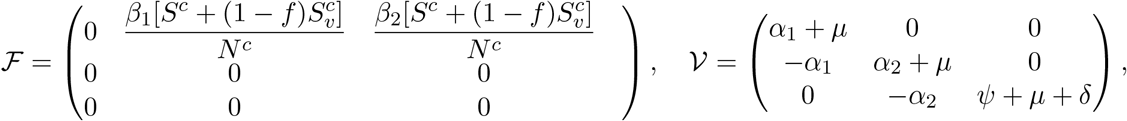

The next generation matrix is defined by ℱ𝒱^*-*1^. Computing the spectral radius (dominant eigenvalue) of the next generation matrix, we obtain the control reproduction number

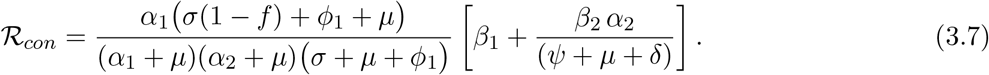

The first term in the control reproduction number (3.7) accounts for the infections caused by the pre-symptomatic individuals in *E*_2_, while the second term account for those caused by symptomatic individuals in (*I*). Using (3.7), we establish the following result.

#### Theorem 3.1.

*The DFE of the COVID-19 model in* (2.2)*-*(2.7) *is locally asymptotically stable (LAS) if ℛ*_*con*_ *<* 1 *and unstable if ℛ*_*con*_ > 1.

The proof of this theorem is standard and can be established using theorem 2 of [59]. Recall that the parameter ℛ_*con*_ is used to measure the average number of new COVID-19 cases generated by a single infected individual introduced into a susceptible population where a certain fraction of individuals are vaccinated. The biological interpretation of Theorem 3.1 is that if the control reproduction number ℛ_*con*_ *<* 1, the influx of a new COVID-19 case in the community will not overwhelm the community but the whole community will be endemic if ℛ_*con*_ > 1.

In Figure 2, we present numerical simulations of the ODE system (2.2)-(2.7) showing the disease-free dynamics. The initial conditions used for this simulation is given in Table 4 and the parameters are given in Table 2. The vaccination rate is set as *σ* = 0.85 with vaccine efficacy *f* = 0.85, while the waning rate for both vaccine-induced and post-recovery immunity are *ϕ*_1_ = *ϕ*_2_ = 0.3. For these set of parameters, the corresponding control reproduction number is ℛ_*con*_ = 0.9896, which is less than 1. As observed from the results in the right panel of this figure, the dynamics of the infected compartments (*E*_1_, *E*_2_, and *I*) tends to zero while the susceptible populations (*S* and *S*_*v*_) initially decrease and then stabilize at some positive numbers (see left panel), which leads to the disease-free equilibrium (DFE), as stated in Theorem 3.1. Using the same set of parameters but increasing the immunity waning rates to *ϕ*_1_ = *ϕ*_2_ = 0.5, the corresponding control reproduction number increases to ℛ_*con*_ = 1.2322. The results for this scenario are shown in Figure 3.As stated by Theorem 3.1, the DFE is unstable for this scenario since ℛ_*con*_ > 1. From the right panel of Figure 3, we observe that unlike in Figure 2, the dynamics of the infected compartments do not tend to zero, rather they saturate as some positive numbers. Similar behaviours can be seen in the inset of the plot in the left panel of Figure 3 for the susceptible populations (*S* and *S*_*v*_) as they reduced drastically but don’t tend to zero. This result shows that the DFE is unable when ℛ_*con*_ > 1, and as a result, the disease cannot be eradicated from the population.

**Figure 2:**
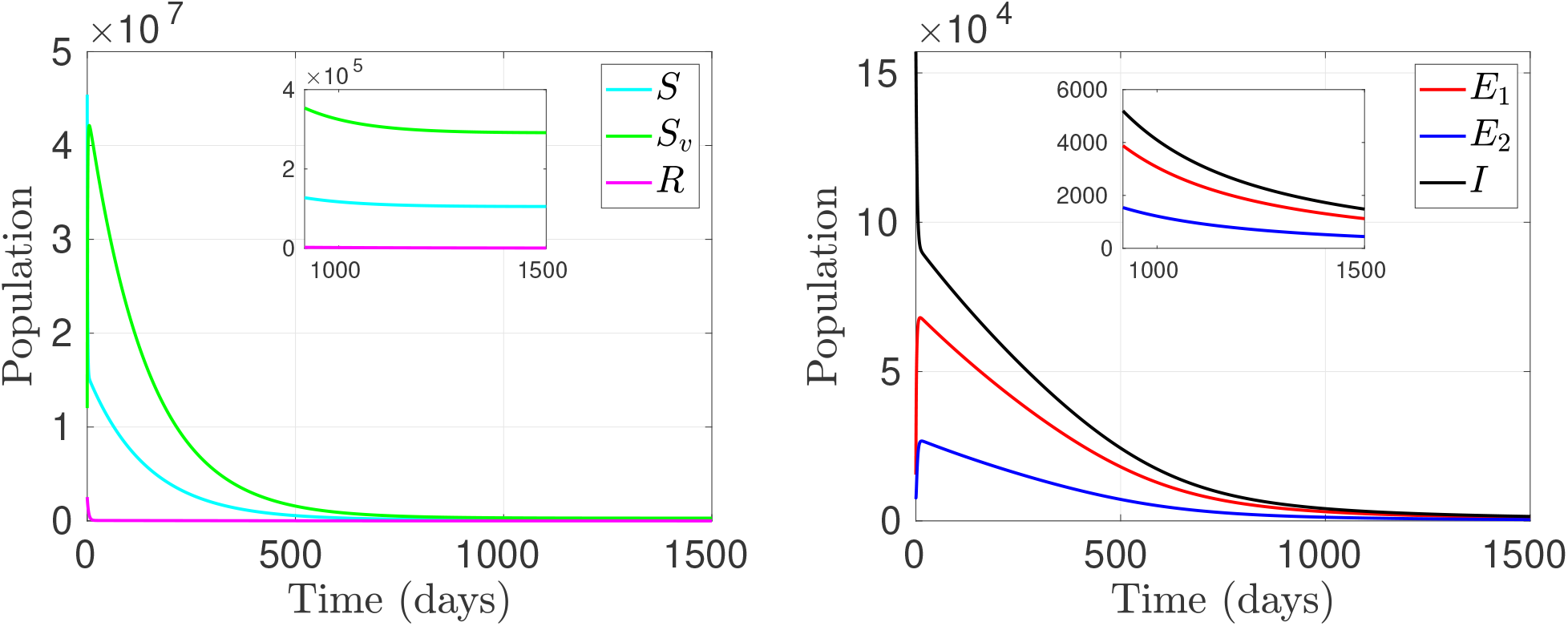
Disease-free dynamics. The time dynamics of the susceptible (S), susceptible-vaccinated (S_v_) and recovered (R) populations (left panel), and those for the exposed (E_1_), pre-symptomatic (E_2_), and symptomatic (I) populations (right panel), showing the disease-free equilibrium dynamics. Vaccination rate is σ = 0.85, with vaccine efficacy, f = 0.85. The vaccine and post-recovery immunity waning rates are ϕ_1_ = ϕ_2_ = 0.3, corresponding to a control reproduction number of ℛ_con_ = 0.9896. Remaining parameters are as given in Table 2.

**Figure 3:**
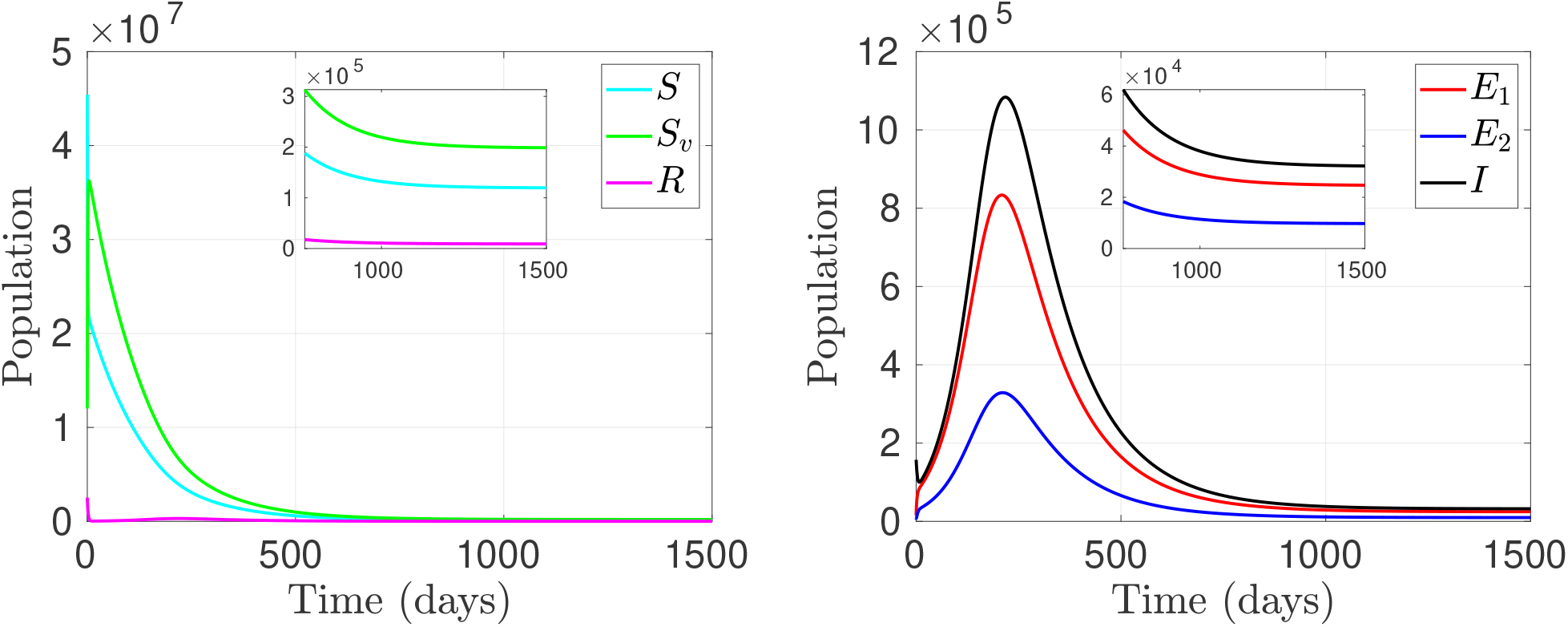
Endemic dynamics. The time dynamics of the susceptible (S), susceptible-vaccinated (S_v_) and recovered (R) populations (left panel), and those for the exposed (E_1_), pre-symptomatic (E_2_), and symptomatic (I) populations (right panel), for when the disease is endemic in the population. Vaccination rate is σ = 0.85, with vaccine efficacy, f = 0.85. The vaccine and post-recovery immunity waning rates are ϕ_1_ = ϕ_2_ = 0.5, corresponding to a control reproduction number of ℛ_con_ = 1.2322. Remaining parameters are given in Table 2.

In the next subsection, we analyze the endemic equilibrium and show that our model undergoes the phe-nomenon of backward bifurcation.

### 3.3 The Backward Bifurcation and Endemic Equilibrium (EE)

Bifurcation is the change in the qualitative structure of the flow of a differential equation with respect to parameter values. The point where bifurcation occurs is called the bifurcation point. Examples of bifurcations in dynamical systems include transcritical, pitchfork, Hopf, saddle-node, forward, backward, e.t.c. [3, 20, 52]. However, the bifurcation type that is related to our study is the backward bifurcation, which is explained in [5] based on the provision of the center manifold theory.

The COVID-19 model presented in (2.2)-(2.7) has a positive and unique endemic equilibrium point, which can be interpreted as the point where at least one infected population exists and is non-zero. Let

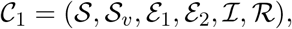

be the endemic equilibrium point. We seek to solve the ODE system (2.2)-(2.7) in terms of the parameter *λ*_*c*_ as follows: The force of infection at steady state can be written as

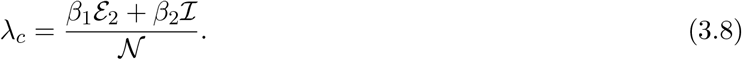

The model equations (2.2)-(2.7) can be re-expressed as

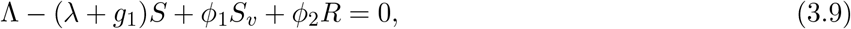

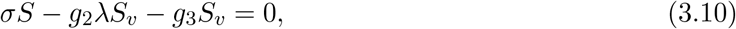

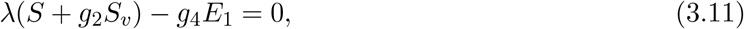

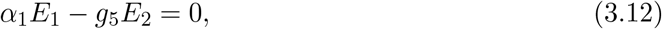

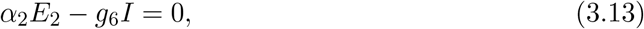

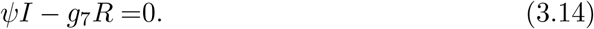

where the newly defined parameters *g*_1_, *g*_2_, …, *g*_7_ are

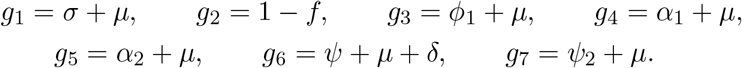

Solving (3.9)-(3.14) simultaneously gives

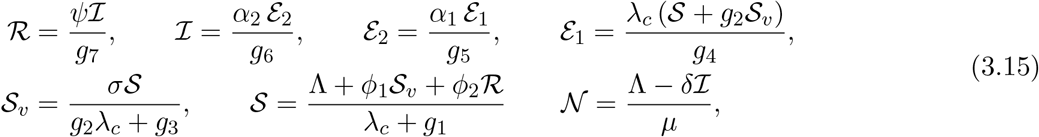

Again, solving the equations in (3.15) simultaneously gives

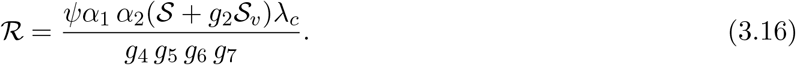

Substituting the expression for *S*_*v*_ in (3.15) into (3.16) and factorizing, we have

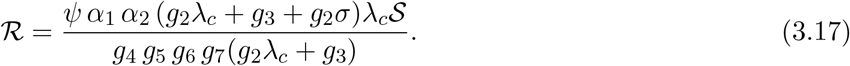

Similarly, we get

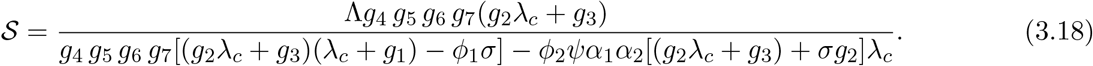

Substituting (3.18) into (3.17) and factorizing gives

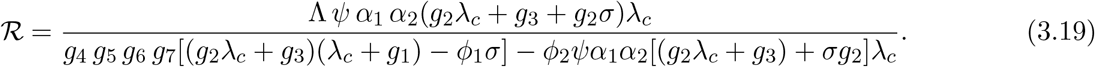

Similarly, we have

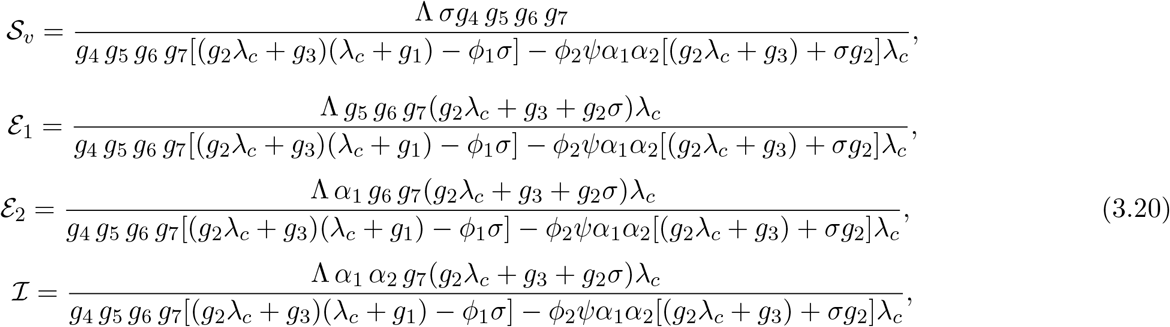

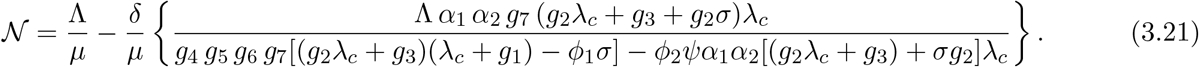

Substituting all the expressions in (3.18), (3.19), (3.20) and (3.21) into (3.8), it can be proved that the non-zero equilibria of the model satisfies

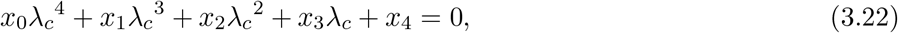

where

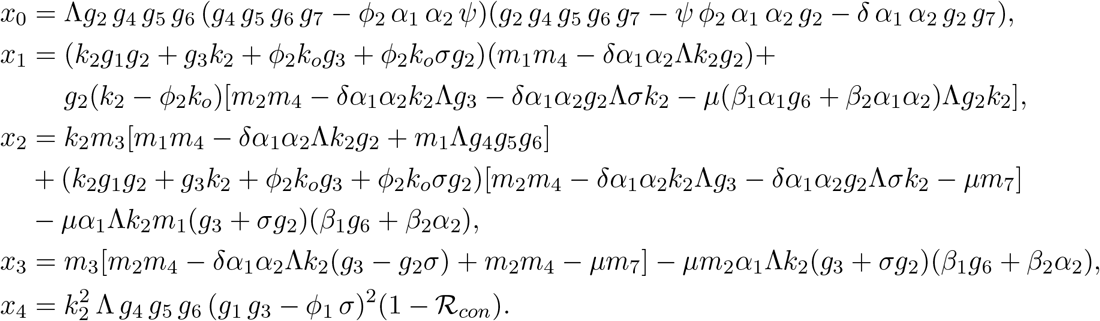

and

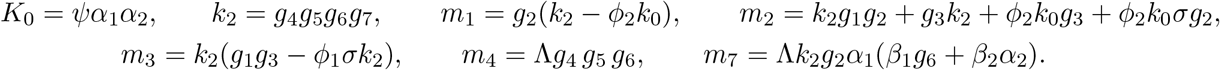

We can easily establish the positiveness of the coefficient *x*_0_ by substituting the respective value of all the parameters in *x*_0_ and subject the resultant expression into full expansion. The negative signs will disappear and the expression will become positive. The complete expansion is cumbersome and thus ignored. More-so, *x*_4_ is positive if and only if ℛ_*con*_ *<* 1 and negative if and only if ℛ_*con*_ > 1. This shows that the number of positive real roots of (3.22) is dependent on the signs of *x*_1_, *x*_2_ and *x*_3_. These signs can be analyzed using the Decartes Rule of signs on (3.22). The possible roots of equation (3.22) are given in Table 3. Hence, we have the following theorem.

**Table 3:** Possible positive real roots of equation (3.22) for ℛ_con_ < 1 and ℛ_con_ > 1.

| Cases | $A_1$ | $A_2$ | $A_3$ | $A_4$ | $A_5$ | $R_0$ | No of times<br>sign changed | No of positive real roots<br>(endemic equilibrium point) |
| --- | --- | --- | --- | --- | --- | --- | --- | --- |
| 1 | + | + | + | + | + | $< 1$ | 0 | 0 |
| | + | + | + | + | - | $> 1$ | 1 | 1 |
| 2 | + | + | + | - | + | $< 1$ | 2 | 0,2 |
| | + | + | + | - | - | $> 1$ | 1 | 1 |
| 3 | + | + | - | + | + | $< 1$ | 2 | 0,2 |
| | + | + | - | + | - | $> 1$ | 3 | 1,3 |
| 4 | + | - | + | + | + | $< 1$ | 2 | 0,2 |
| | + | - | + | + | - | $> 1$ | 3 | 1,3 |
| 5 | + | - | - | + | + | $< 1$ | 2 | 0,2 |
| | + | - | - | + | - | $> 1$ | 3 | 1,3 |
| 6 | + | - | - | - | + | $< 1$ | 2 | 0,2 |
| | + | - | - | - | - | $> 1$ | 1 | 1 |
| 7 | + | - | + | - | + | $< 1$ | 4 | 0,2,4 |
| | + | - | + | - | - | $> 1$ | 3 | 1,3 |
| 8 | + | + | - | - | + | $< 1$ | 2 | 0,2 |
| | + | + | - | - | - | $> 1$ | 1 | 1 |

#### Theorem 3.2.

*The COVID-19 model* (2.2)*-*(2.7)

1. *has a unique endemic equilibrium if ℛ*_*con*_ > 1 *and whenever cases 1, 2, 6 and 8 of Table 3 are satisfied;*
2. *could have more than one endemic equilibrium if ℛ*_*con*_ > 1 *and cases 3, 4, 5 and 7 of Table 3 are satisfied;*
3. *could have 2 or more endemic equilibria if R*_*con*_ *<* 1 *and cases 2-8 of Table 3 are satisfied*.

#### Lemma 2.

*The COVID-19 model has at least one positive endemic equilibrium whenever ℛ*_*con*_ > 1 *but could have zero, two or four positive endemic equilibrium whenever ℛ*_*con*_ *<* 1.

It is worth noting that the exact expression for the endemic equilibria can be obtained by solving for *λ*_*c*_ in (3.22) to obtain the roots and then substitute its positive value into the expressions in (3.18), (3.19), (3.20) and (3.21). The existence of multiple endemic equilibria when the control reproduction number is less than unity established using Theorem 3.2, Lemma 2 and Table 3 suggests the possibility of backward bifurcation [3, 52]. As previously mentioned, backward bifurcation is a phenomenon where a stable DFE coexists with a stable endemic equilibrium when the control reproduction number is less than 1. We present the bifurcation diagrams in Figure 4.

**Figure 4:**
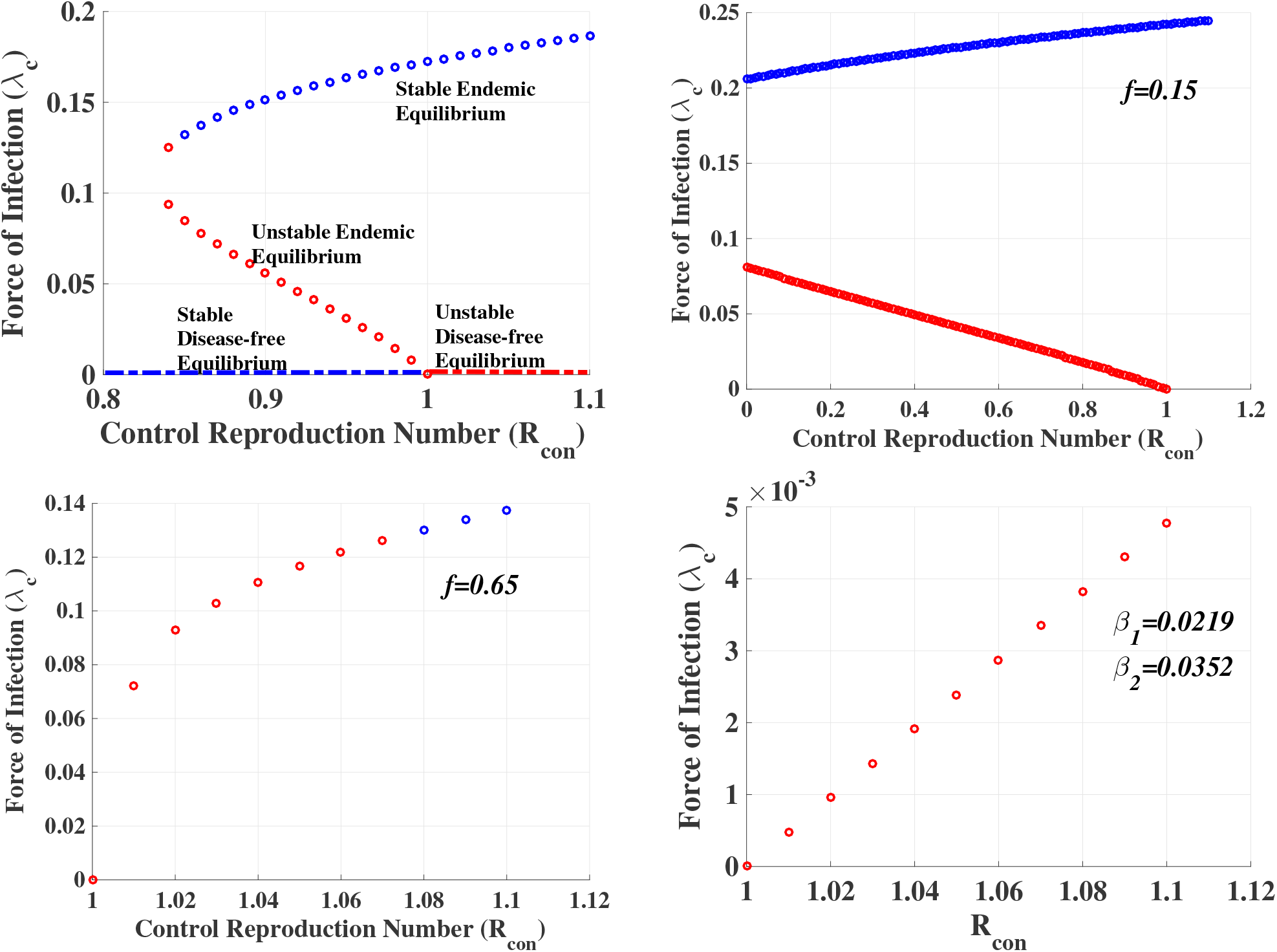
Bifurcation Diagrams. The bifurcation diagrams for different values of vaccine efficacy f and different transmission rates β_1_ and β_2_. The top-left panel is the original bifurcation diagram for f = 0.55 and σ = ϕ_1_ = ϕ_2_ = 0.5 while other parameter values remain unchanged. The top-right panel is the bifurcation diagram for f = 0.15 and σ = ϕ_1_ = ϕ_2_ = 0.5 while other parameter values remain unchanged. The bottom-left panel shows the bifurcation diagram for f = 0.65 and σ = ϕ_1_ = ϕ_2_ = 0.5 while other parameter values remain unchanged. The bottom-right panel shows the bifurcation diagram f = 0.55, σ = ϕ_1_ = ϕ_2_ = 0.5 and β_1_ = 0.0219, β_2_ = 0.0352 (which is 10% of its baseline value) while other parameter values remain unchanged.

The bifurcation diagram on the top left panel of Figure 4 shows the behaviour of the endemic model (2.2)-(2.7) when the control reproduction number ℛ_*con*_ is less than, equal and greater than 1. The parameter values used are given in Table 1 with *σ* = *ϕ*_1_ = *ϕ*_2_ = 0.5 and vaccine efficacy set at *f* = 0.55. The red-circled line on the diagram shows that the endemic equilibrium is unstable for ℛ_*con*_ *<* 1 while the blue-circled part of the diagram shows the stability of the endemic equilibrium for ℛ_*con*_ > 1. The blue-dashed line shows the stability of the disease-free equilibrium for ℛ_*con*_ *<* 1 while the red-dashed line shows that the disease-free equilibrium is unstable for *R*_*con*_ > 1. We observe from this result that there are two steady-states of the system (DFE and endemic equilibrium) when ℛ_*con*_ > 1. As ℛ_*con*_ decreases, we have the emergence of an endemic equilibrium (red-circled line) at ℛ_*con*_ = 1, which is unstable, that co-exist with a stable endemic equilibrium (blue-circled line) and a stable disease-free equilibrium (blue-dashed line). The backward movement of the equilibrium that emerge at ℛ_*con*_ = 1 clearly shows that our model exhibits the phenomenon of backward bifurcation, where a stable disease-free equilibrium point co-exists with a stable endemic equilibrium point. The epidemiological implication of this is that the control reproduction number being less than unity is no longer sufficient to guarantee the disease eradication. More control measures would be required to eradicate the disease. For this reason, we considered the effects of increase in vaccine efficacy and reduction in transmission rates on the bifurcation diagram.

The top right panel shows the behavior of the bifurcation diagram when the vaccine efficacy *f* is reduced to 0.15 while other parameter values remain the same. We can see from this result that bi-stability persist, and now exist for all ℛ_*con*_ *<* 1. The bottom left panel shows the behavior of the model when the vaccine efficacy *f* is increased to 0.65 while other parameter values remain the same. We can easily notice the disappearance of bi-stability. Also, we observe that the endemic equilibrium is unstable for some values of ℛ_*con*_ > 1. For these values of ℛ_*con*_, the DFE equilibrium is stable (not shown). This clearly underlines the need for a highly efficacious vaccine in controlling the spread of the disease and its eventual eradication. The bottom right panel shows the behaviour of the model when the transmission rates *β*_1_ and *β*_2_ are reduced to 0.02189 and 0.03521, respectively (which is 10% of its baseline value) while other parameter values remain the same. We observe from this result that there is also no bi-stability in the system for this scenario. This result emphasizes the importance of reduction in community transmission in the fight against the disease. The lower/higher the transmission rates the lower/higher the disease prevalence. On the other hand the higher/lower the vaccine efficacy the higher/lower the protection against the disease. These two parameters are important in controlling the disease spread and its eventual eradication.

## 4 Numerical simulations

We present the numerical simulations of the endemic model (2.2)-(2.7) for different scenarios to study the effect of immunity waning on the dynamics of COVID-19. The parameters used are related to the demography of South Africa. These parameters are given in Table 2. We studied the effect of immunity waning on the control reproduction number and the entire diseases dynamics.

In Figure 5, we present the contour plots of the control reproduction number ℛ_con_ in terms of the vaccine immunity waning parameter *ϕ*_1_ and the vaccination rate *σ* for vaccine efficacy *f* = 0.5 (left panel) and *f* = 0.85 (right panel). We observe from the results in this figure that the control reproduction number, ℛ_con_ decreases as the vaccination rate increases and increases as the vaccine immunity waning rates increase. When the vaccination rate is relatively small, vaccine immunity waning has little effect on the reproduction number, compare to when a large fraction of the population is vaccinated. In this case, an increase in the waning rate increases the control reproduction number. In addition, comparing the results in the left panel of this figure for vaccine efficacy *f* = 0.5 to that of the right panel for *f* = 0.85, we notice that the vaccine immunity waning rate has more effect on the control reproduction number when the vaccine is more effective.

**Figure 5:**
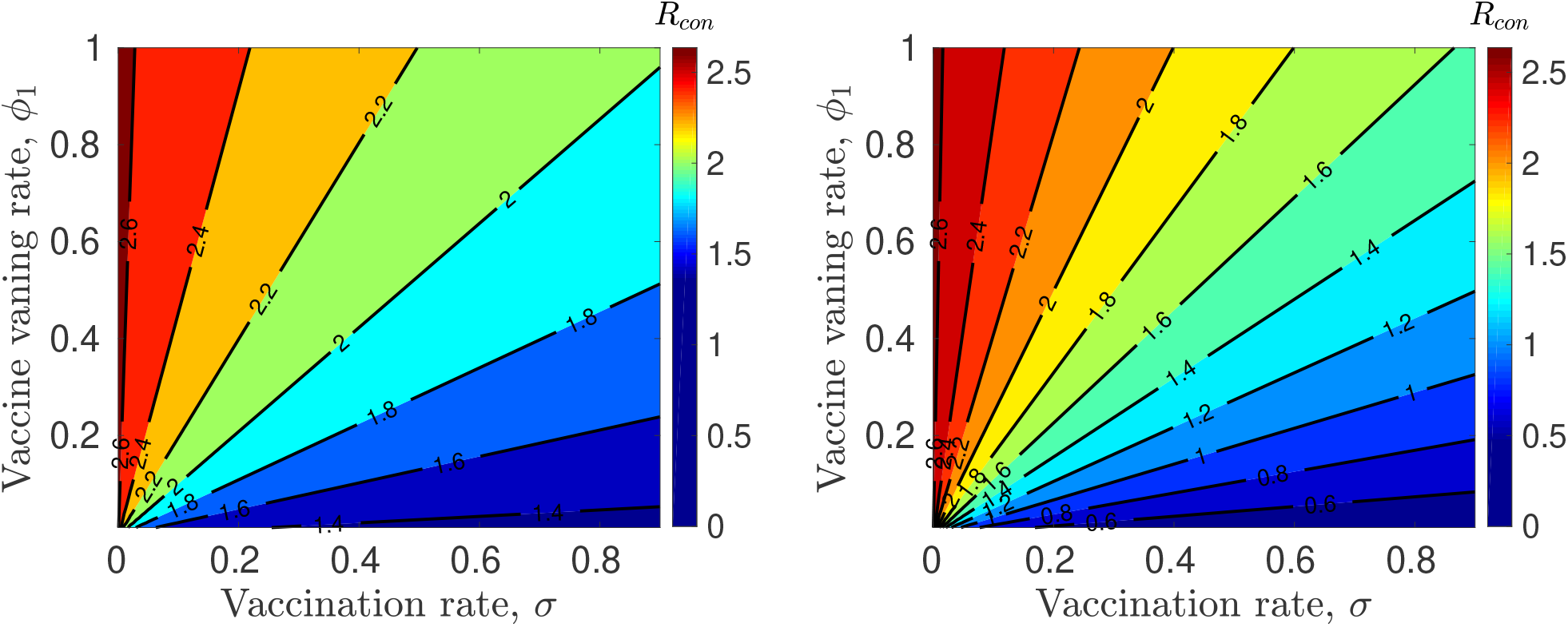
Effect of vaccination and vaccine immunity waning on the control reproduction number. Left panel: vaccine efficacy, f = 0.5. Right panel: vaccine efficacy, f = 0.85. Remaining parameters are given in Table 2.

Similar results are shown in Figure 6 for the transmission and vaccination rates. In the top left panel of this figure, we show the control reproduction number as a function of the pre-symptomatic transmission rate (*β*_1_) and the vaccination rate (*σ*) for *ϕ*_1_ = 0.1. For the same vaccine waning rate, we present the reproduction number as a function of the symptomatic transmission rate (*β*_2_) and the vaccination rate (*σ*).

**Figure 6:**
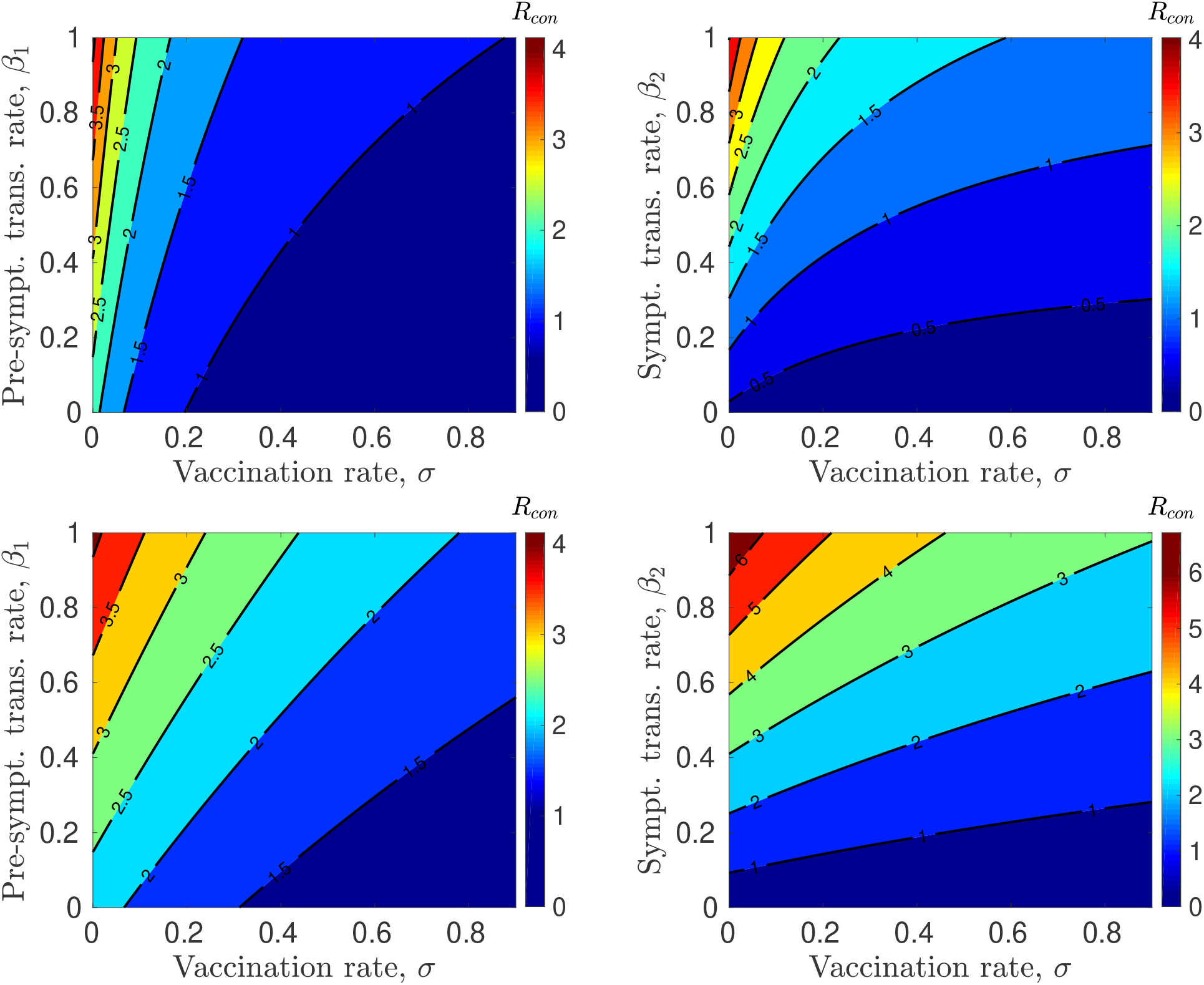
Effect of vaccination rate and disease transmission rates on the control reproduction number. Left panel: Pre-symptomatic transmission rate β_1_, and right panel: Symptomatic transmission rate β_2_. In the first row, the vaccine immunity waning rate is ϕ_1_ = 0.1, and ϕ_1_ = 0.5 in the second row. Vaccine efficacy is set as f = 0.85. Remaining parameters are given in Table 2.

The remaining parameters are given in Table 2. As expected, the control reproduction number increases as the transmission rates *β*_1_ and *β*_2_ increase, although, the symptomatic transmission rate has more effect on the control reproduction number compared to the pre-symptomatic transmission rate. The results in the lower panel of Figure 6 are similar to those in the top panel but for a higher waning rate of vaccine-induced immunity (*ϕ*_1_ = 0.5). Comparing these results with those in the top panel, we notice that an increase in *ϕ*_1_ increases the dependence of the control reproduction number on the vaccination rate. Similarly, when *ϕ*_1_ is small (say *ϕ*_1_ = 0.1 as shown in the top panel of Figure 6), the transmission rates have more effect on the reproduction number when a small fraction of the population is vaccinated compared to when the vaccination rate is high. However, the effect of the transmission rates on the control reproduction number when vaccination rate is higher increases as the immunity waning rate increases. This can easily be seen by comparing the results in the top panel to those in the bottom panel for high vaccine efficacy.

Next, we study the effect of immunity waning on the dynamics of COVID-19 using South Africa as a case study. The results of this study are presented in terms of the time dynamics of the symptomatic infectious population. The initial conditions used are presented in Table 4 and are based on the South African COVID-19 statistics as of August 29, 2021, and were obtained from the Department of health COVID-19 report, Republic of South Africa [11]. In addition, the parameters used are also related to the demography of South Africa and are presented in Table 2. Figure 7 shows the dynamics of the symptomatic population over time for different vaccine immunity waning rate, *ϕ*_1_ (left panel) and post-recovery immunity waning rate, *ϕ*_2_ (right panel). We observe from these results that vaccine immunity waning has much more effect on the disease dynamics than the post-recovery immunity waning. For this scenario, we used *ϕ*_1_ = 0.3, 0.5, 0.75, and 0.9, which give the corresponding control reproduction numbers ℛ_*con*_ = 0.9896, 1.2322, 1.4504, and 1.5516, respectively. By varying the vaccine waning rate from *ϕ*_1_ = 0.3 through *ϕ*_1_ = 0.9, the control reproduction number goes from ℛ_*con*_ = 0.9896 (ℛ_*con*_ *<* 1) to ℛ_*con*_ = 1.5516 (ℛ_*con*_ > 1). Changing the disease dynamics from disease-free to endemic (See the left panel of Figure 7). However, this is not the case for the post-recovery immunity waning rate *ϕ*_2_. This parameter has little effect on the dynamics of the disease (right panel of Figure 7). In addition, since this control reproduction number is independent of this parameter, ℛ_*con*_ = 1.2322 for all values of *ϕ*_2_.

**Table 4:** Model initial conditions. These initial conditions are based on the South African COVID-19 statistics as of August 29, 2021, and were obtained from the Department of health COVID-19 report, Republic of South Africa [11]. The unvaccinated susceptible population was computed using S(0) = N (0) − (S_v_(0) + E_1_(0) + E_2_(0) + I(0) + R(0)).

| Variable | Initial population | Value | References |
| --- | --- | --- | --- |
| $N(0)$ | Total population | 60,142,978 | [9] |
| $S(0)$ | Unvaccinated susceptible population | 45,414,814 | Computed |
| $S_v(0)$ | Vaccinated susceptible population | 12,021,608 | [10, 11] |
| $E_1(0)$ | Exposed population | 15,480 | Inferred [11] |
| $E_2(0)$ | Pre-symptomatic population | 7,740 | Inferred [11] |
| $I(0)$ | Symptomatic population | 157,137 | [11] |
| $R(0)$ | Recovered population | 2,526,199 | [11] |

**Figure 7:**
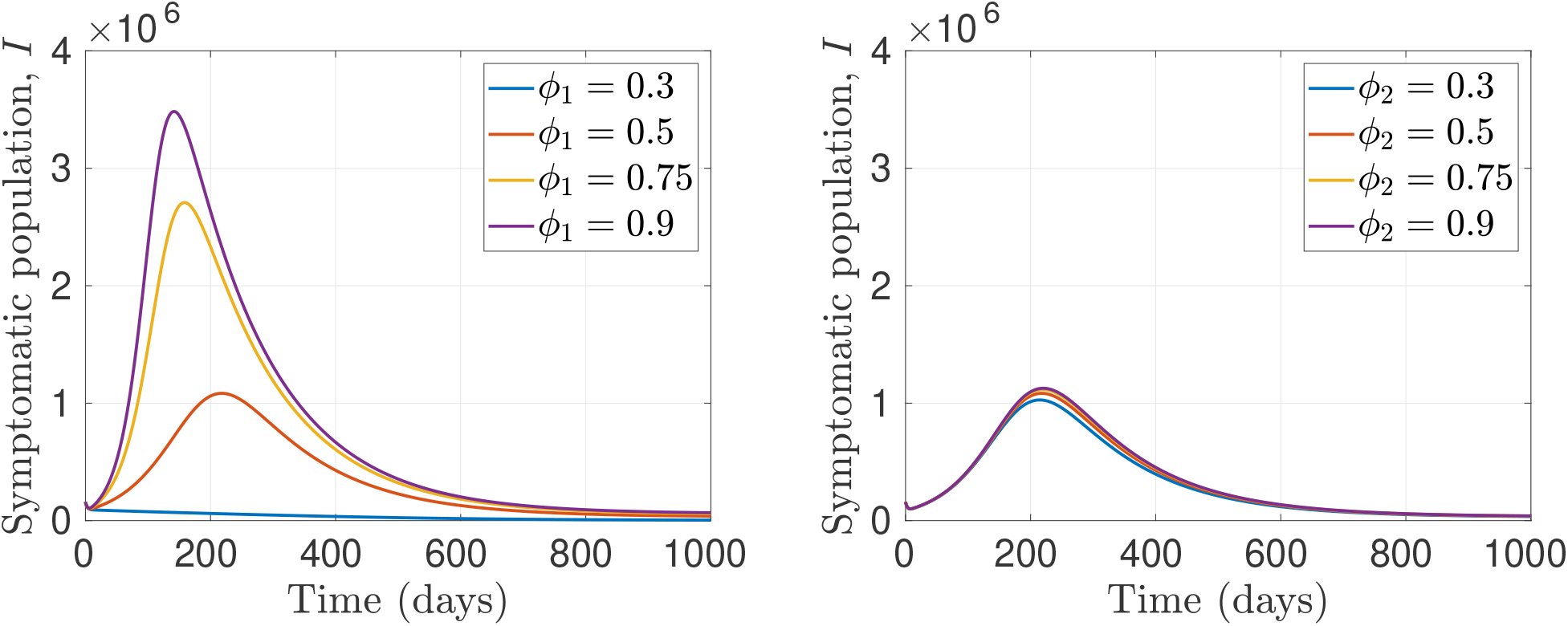
Effect of immunity waning on disease dynamics. The dynamics of the symptomatic population over time for different vaccine immunity waning rates (left panel) and post-recovery immunity waning rates (right panel). In the left panel, for the waning rates ϕ_1_ = 0.3, 0.5, 0.75, and 0.9, the corresponding control reproduction numbers are ℛ_con_ = 0.9896, 1.2322, 1.4504, and 1.5516, respectively, while in the right panel, ℛ_con_ = 1.2322 for all values of ϕ_2_. Vaccination rate used is σ = 0.85, with vaccine efficacy, f = 0.85. Remaining parameters are given in Table 2.

In Figure 8, we study the effect of the vaccination rate, *σ* and the immunity waning rates *ϕ*_1_ and *ϕ*_2_ on the disease dynamics. In this figure, we reduce the vaccination rate, from *σ* = 0.85 used in Figure 7 to *σ* = 0.5. Our goal is to study how a reduction in vaccination rate would affect the disease dynamics. Comparing the results in Figure 7 to those in Figure 8, we observe that a decrease in vaccination rate lead to more infections in the population. Unlike the case where vaccination rate is high, when vaccination rate is *σ* = 0.5, there are no disease-free dynamics for all values of the waning parameter *ϕ*_1_ used. This results suggests that we need to keep the vaccination rate high in order to eradicate the disease.

**Figure 8:**
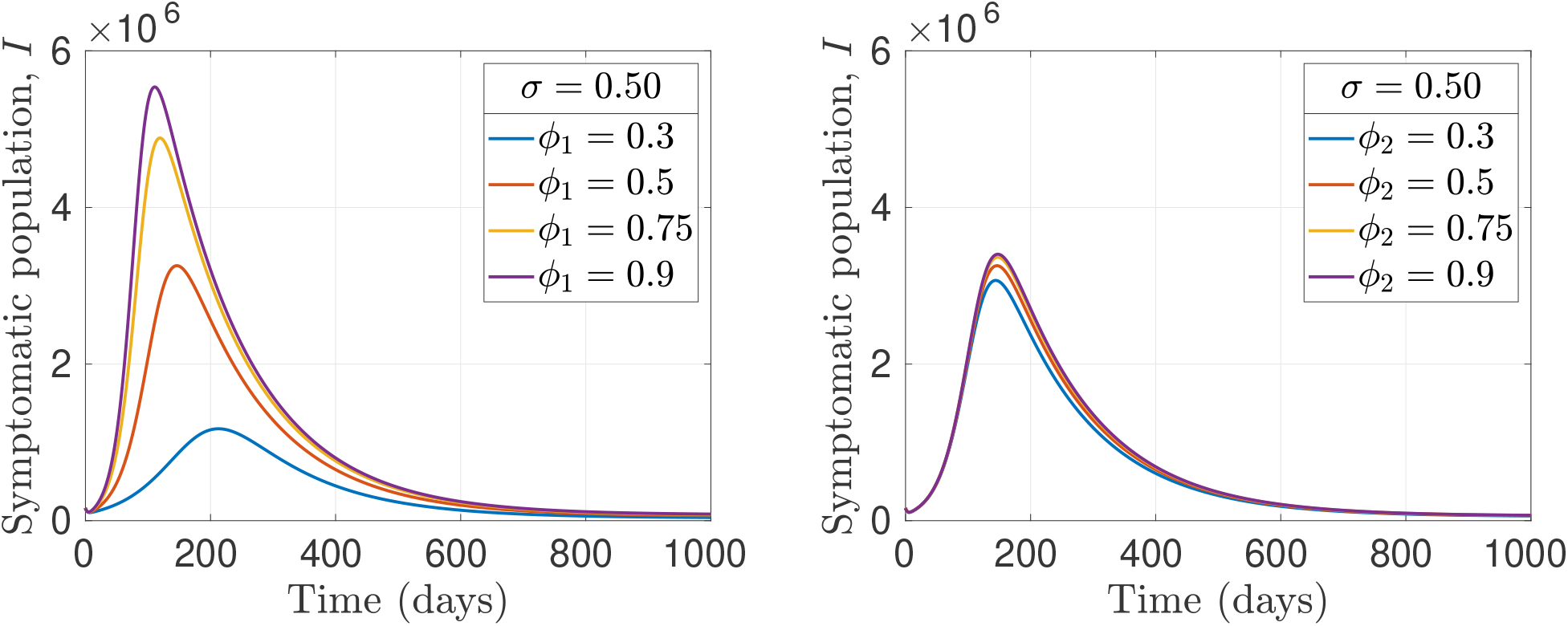
Effect of vaccination rate and immunity waning on disease dynamics. The dynamics of symptomatic population over time for different vaccine immunity waning rates (left panel) and post-recovery immunity waning rates (right panel). In the left panel, for the waning rates ϕ_1_ = 0.3, 0.5, 0.75, and 0.9, the corresponding control reproduction numbers are ℛ_con_ = 1.2474, 1.5230, 1.7442, and 1.8392, respectively, while in the right panel, ℛ_con_ = 1.5230 for all values of ϕ_2_. Vaccination rate is reduced to σ = 0.5, with vaccine efficacy, f = 0.85. Remaining parameters are given in Table 2.

The result in Figure 9 is for when the vaccine efficacy is reduced from *f* = 0.85 used in Figure 7 to *f* = 0.65, while the vaccination rate remains *σ* = 0.85. Similar to the results in Figure 8, the disease is endemic in the population for all values of the waning parameter *ϕ*_1_. The increase in infections observed in Figure 9 relative to Figure 7 is due to a decrease in the efficacy of the vaccine, since the same vaccination rate is used for the two scenarios. This suggests that in addition to high vaccination rate, a high vaccine efficacy is also essential to eradicate the disease.

**Figure 9:**
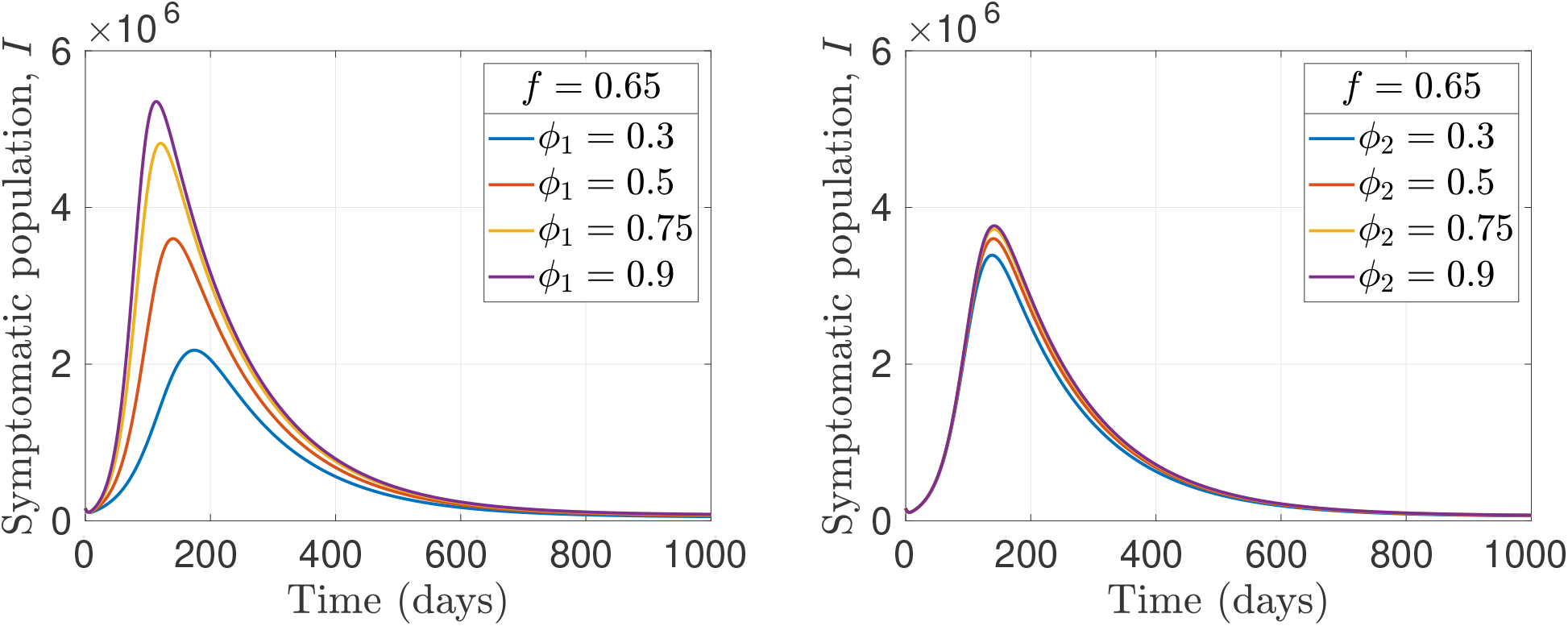
Effect of vaccine efficacy and immunity waning on disease dynamics. The dynamics of symptomatic population over time for different vaccine immunity waning rates (left panel) and post-recovery immunity waning rates (right panel). In the left panel, for the waning rates ϕ_1_ = 0.3, 0.5, 0.75, and 0.9, the corresponding control reproduction numbers are ℛ_con_ = 1.3768, 1.5623, 1.7292, and 1.8065, respectively, while in the right panel, ℛ_con_ = 1.5623 for all values of ϕ_2_. Vaccination rate is σ = 0.85, with vaccine efficacy reduced to f = 0.65. Remaining parameters are given in Table 2.

Lastly, we study the effect of the transmission rates on the disease dynamics. In the top panel of Figure 10, we present the results for when the pre-symptomatic transmission rate is increased to *β*_1_ = 0.3284, while the symptomatic transmission rate remains the same as in the remaining figures (*β*_2_ = 0.3521). On the other hands, in the lower panel we increased the symptomatic transmission rate to *β*_2_ = 0.4049, while the pre-symptomatic transmission rate remains the same as in the other figures (*β*_1_ = 0.2189). As expected, there are more infections are the immunity waning rates increase. Comparing the results in this figure to those in Figure 7, we observe that an increase in the transmission rates will increase the disease spread in the community making it difficult to eradicate the disease. In addition, an increase in the symptomatic transmission rate significantly affect the disease dynamics relative to the pre-symptomatic transmission rate. Note that for all the examples considered, the post-recovery immunity waning rate has little effect on the disease dynamics compared to the waning rate of vaccine-induced immunity.

**Figure 10:**
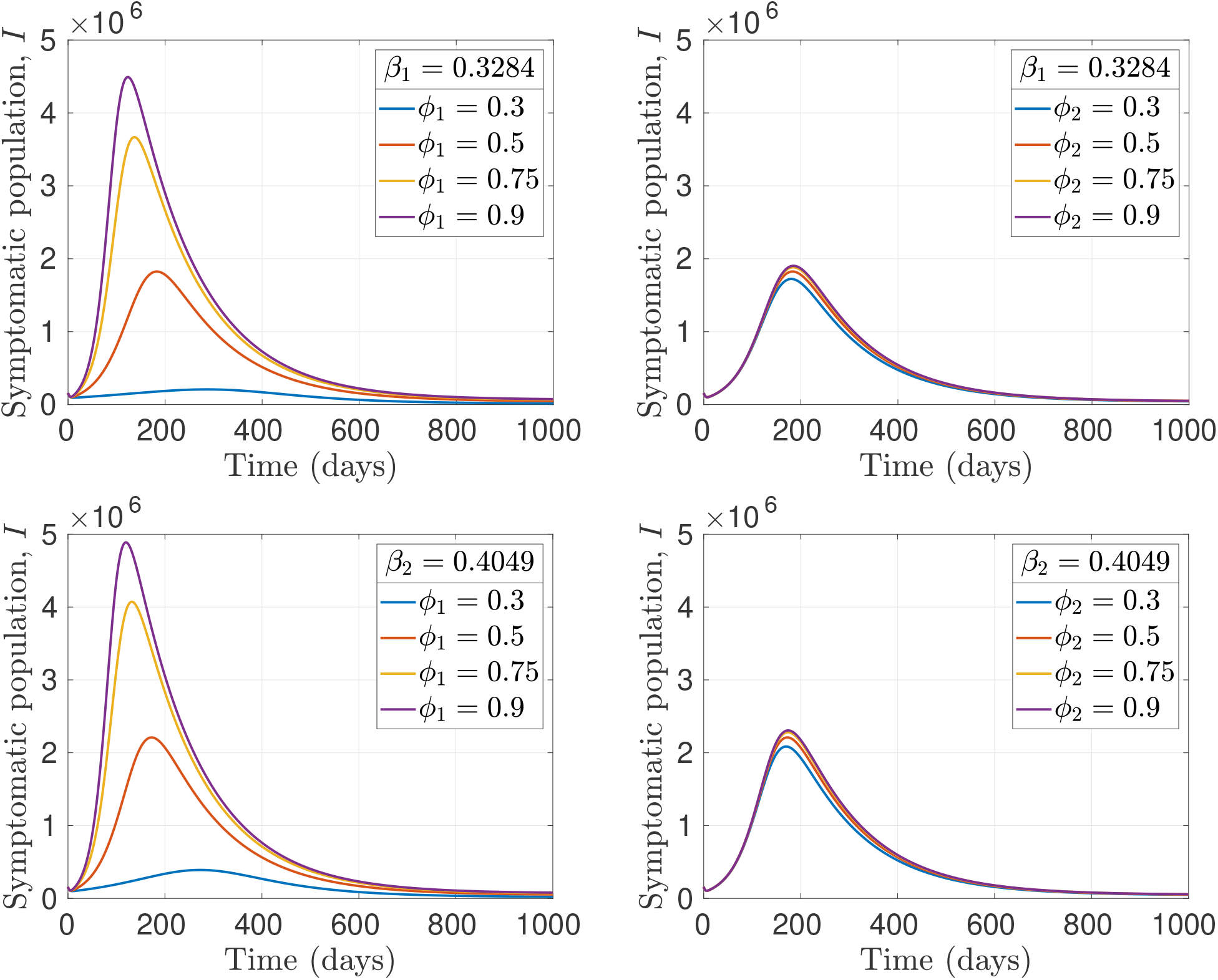
Effects of disease transmission rates and immunity waning on disease dynamics. The dynamics of symptomatic population over time for different vaccine immunity waning rates (left) and post-recovery immunity waning rates (right). In the top panel, the pre-symptomatic transmission rate is β_1_ = 0.3284 and the symptomatic transmission rate is β_2_ = 0.3521, while in the lower panel, the pre-symptomatic transmission rate is β_1_ = 0.2189 and the symptomatic transmission rate is β_2_ = 0.4049. In the top left panel, for the waning rates ϕ_1_ = 0.3, 0.5, 0.75, and 0.9, the corresponding control reproduction numbers are ℛ_con_ = 1.0680, 1.3297, 1.5652, and 1.6744, respectively, while in the top right panel, R_con_ = 1.5623 for all values of ϕ_2_. Similarly, in the bottom left panel, for the waning rates ϕ_1_ = 0.3, 0.5, 0.75, and 0.9, the corresponding control reproduction numbers are ℛ_con_ = 1.1146, 1.3877, 1.6336, and 1.7475, respectively, while in the bottom right panel, ℛ_con_ = 1.3877 for all values of ϕ_2_. Remaining parameters are given in Table 2.

## 5 Discussion

We have developed and analyzed an endemic model for COVID-19 to study the effect of immunity waning on the dynamics of thes disease. Our model has six compartment: unvaccinated susceptible (*S*), vaccinated susceptible (*S*_*v*_), exposed (not infectious) (*E*_1_), pre-symptomatic (infectious but no symptoms), symptomatic (*I*), and recoversed (*R*). We considered two type of immunity waning in our model: vaccine immunity waning and post-recovery immunity waning. Vaccine immunity waning occurs when a vaccinated individual in the *S*_*v*_ compartment loses their immunity and return to the unvaccinated susceptible compartment *S*. On the other hand, post-recovery immunity waning occurs when recovered individuals lose their immunity and return to the unvaccinated susceptible compartment. Both immunity waning described above can also be interpreted as being susceptible to another strain of COVID-19. Our model implements a sterilizing vaccine that reduces the chances of getting infected. We apply our model to the South African COVID-19 scenario.

We analyzed our model rigorously by first computing the control reproduction number for the disease-free equilibrium. After which we analyzed the endemic equilibrium and show that our model exhibit the backward bifurcation phenomenon. We examined the condition for the existence of bifurcation and endemic equilibrium. These conditions are stated in Theorem 3.2. Furthermore, Lemma 2 provides the condition for the existence of multiple endemic equilibria with respect to the conditions specified on Table 3. This leads to the phenomenon where stable DFE co-exists with a stable endemic equilibrium when the control reproduction number is less than unity. Figure 4 shows the bifurcation diagrams for different scenarios. It can be understood from this diagrams that if the vaccine efficacy is reduced, bi-stability persist but when increased, the bi-stability disappears and the disease may die out. A better result is achieved when the transmission rates are reduced, thereby eliminating the bi-stability totally. This clearly underlines the need for a highly efficacious vaccine and reduction in disease transmission to achieve disease eradication.

We numerically studied the effects of vaccination and the waning of vaccine immunity on the control reproduction number. These results were presented in form of contour plots and show that the control reproduction number decreases as the vaccination rate increases. However, it increases as the vaccine immunity waning rate increases. As the vaccine efficacy increases, the effects these parameters (vaccination rate and vaccine immunity waning rate) become more pronounced. There is also a decrease in the control reproduction number, although, the decrease is more when the vaccination rate is high. When a small fraction of the population is vaccinated, vaccine immunity waning has little effect on the control reproduction number, compared to when vaccination rate is high. This is because when a large fraction of the population is vaccinated, we expect a reduction in infection. But when these people loose their immunity at a faster rate, the expected reduction in infection will not be realized, leading to a significant spread of the disease. This is not the case when only a small fraction of the population are vaccinated from the onset. In this case, there will be no much difference whether they loose their immunity slowly or at a faster rate. We also studied the effect of the transmission rates together with vaccination on the reproduction number. Our results show that the control reproduction number increases as the transmission rates increase, although, the symptomatic transmission rate has more effect on the control reproduction compared to the pre-symptomatic transmission rate. This may be because infectious individuals spend fewer days (2 days) in the pre-symptomatic compartment compared to the symptomatic compartment, where they spend 7 days. In addition, the transmission rates have little effect on the reproduction number when vaccination rate is high, and this effect increases as the vaccination rate drops. In other words, when we vaccinate a large fraction of the population with a relatively effective vaccine, the transmission rate has a little effect on the reproduction number. However, if we vaccinate only a small fraction of the population with the same vaccine when the transmission rate is higher, the control reproduction number increases too.

Furthermore, we investigated the effects of different parameters of the model on the disease dynamics. Our main focus is studying how vaccination and the waning of immunity affect the disease dynamics. The results of these investigations were presented in terms of the dynamics of the symptomatic population over time and show that vaccine immunity waning has more effect on the disease dynamics compared to the post-recovery immunity waning. By varying the vaccine immunity waning parameter, we realized that when this parameter is low, the diseased-free equilibrium (DFE) of the model is linearly asymptotically stable. This means that the disease can be eradicated when people retain their vaccine immunity for a longer time. However, as this waning rate is increased, the DFE becomes unstable, there by making the disease endemic in the population. On the other hand, the DFE remains unstable for all values of the post-recovery immunity waning parameter and a moderate vaccine waning parameter. By reducing the vaccination rate by ≈ 40%, we noticed that there is no value of vaccine immunity waning rate for which the DFE is linearly stable. As a result of this, the disease cannot be eradicated when the vaccination rate is low. In summary, keeping the vaccination rate high and preventing lose of immunity is required to eradicate COVID-19.

An interest future direction for this work is to fit our model to the reported cases of COVID-19 for a specific location/country. In this way, the parameters would be more related to that location and the predictions would be more representative of the location. Another interesting future direction is to extend the model to have explicit cycles of infection or loss of immunity. For this case, there will be all six compartments of our current model in each cycle such that individuals that lose their immunity in one cycle will go to the susceptible compartment of the next cycle rather than the susceptible compartment of their current cycle as it is in our current model, and the disease will progress from there to the other compartments of the next cycle. An advantage of structuring the model this way is that one can easily track the number of people getting infected at each infection cycle. In addition, different cycles of infection or loss of immunity can be interpreted as being susceptible to different variants of COVID-19, where different vaccine efficacy could also be used. This will increase the flexibility of the model. It would also be worthwhile to stratify the population by age and extend the model to have age-structure. This is particularly important since individuals younger than 12 years have not been approved for vaccination yet. It would also make the model more suitable for answering age related questions.

Some limitations of the modeling framework presented in this paper includes modeling vaccination as a single dose despite the fact that majority of the COVID-19 vaccines available today require more than one does to get a good level of protection. In addition, our model assumes that vaccine efficacy is the same, either one is getting the vaccine for the first time or one had already experienced waning of immunity. This may not be the case in reality as getting the vaccine the second time may lead to a better/stronger protection. Also the efficacy of the vaccines for different variants of COVID-19 may be different. Another limitation of our model is assuming that the population is well-mixed. This may not be so in reality as contact rates and mixing patterns may vary between individuals depending on their age and activity level. We would also like to include lack of stochasticity as a limitation to our modeling framework. Despite all these limitations, we believe that our modeling framework has provided some insights into the endemic dynamics of COVID-19 and has also provided some reasonable suggestions on the important parameters to be considered to effective eradicate the disease.

## Data Availability

All data produced in the present work are contained in the manuscript

## Acknowledgement

No funding is received for this project.

## Data Availability

All used data are properly referenced in the body of the work.

